# Comparison of the mucosal and systemic antibody responses in Covid-19 recovered patients with one dose of mRNA vaccine and unexposed subjects with three doses of mRNA vaccines

**DOI:** 10.1101/2022.12.16.22283554

**Authors:** Shaojun Liu, Joseph GS Tsun, Genevieve PG Fung, Grace CY Lui, Kathy YY Chan, Paul KS Chan, Renee WY Chan

## Abstract

**Background:** Immunity acquired from natural SARS-CoV-2 infection and vaccine wanes overtime. This longitudinal prospective study compared the effect of a booster vaccine (BNT162b2) in inducing the mucosal (nasal) and serological antibody between Covid-19 recovered patients and healthy unexposed subjects with two dose of mRNA vaccine (vaccine-only group).

**Method:** Eleven recovered patients and eleven gender-and-age matched unexposed subjects who had mRNA vaccines were recruited. The SARS-CoV-2 spike 1 (S1) protein specific IgA, IgG and the ACE2 binding inhibition to the ancestral SARS-CoV-2 and omicron (BA.1) variant receptor binding domain were measured in their nasal epithelial lining fluid and plasma.

**Result:** In the recovered group, the booster expanded the nasal IgA dominancy inherited from natural infection to IgA and IgG. They also had a higher S1-specific nasal and plasma IgA and IgG levels with a better inhibition against the omicron BA.1 variant and ancestral SARS-CoV-2 when compared with vaccine-only subjects. The nasal S1-specific IgA induced by natural infection lasted longer than those induced by vaccines while the plasma antibodies of both groups maintained at a high level for at least 21 weeks after booster.

**Conclusion:** The booster benefited all subjects to obtain neutralizing antibody (NAb) against omicron BA.1 variant in plasma while only the Covid-19 recovered subjects had an extra enrichment in nasal NAb against Omicron BA.1 variant.

## 1. Introduction

Coronavirus disease 2019 (Covid-19) is an infectious disease caused by the severe acute respiratory syndrome coronavirus 2 (SARS-CoV-2). We have lived with the SARS-CoV-2 for more than two and a half years while the immune landscape of the population and the SARS-CoV-2 changes over time. Up to now, over 617 million of Covid-19 cases have been reported wordwide and 67.9% of the world population has received at least one dose SARS-CoV-2 vaccine [1]. With the introduction of new variant of concern (VOC), namely Alpha, Beta, Gamma, Delta and Omicron, identified in the United Kingdom in September 2020, South Africa in May 2020, Brazil in November 2020, India in October 2020 and multiple countries in November 2021, respectively [2], the emergence of the omicron sub-lineages BA.2 since March 2022 and the continuous evolution and upsurge of BA.5 [3], whether a prior infection or the vaccination designed for the original SARS-CoV-2 strain would provide us with sufficient protection against the new VOCs is not guaranteed.

So far, Hong Kong has experienced five waves of Covid-19 outbreaks, with 2.06 million confirmed cases and 10,634 deaths. Since the appearance of the first Covid-19 case in Hong Kong in early 2020, the Department of Health in Hong Kong implemented intense surveillance measures [4] and vigorous contact tracing by the Centre of Health Protection for early quarantine and isolation. It was a very successful strategy to combat the first four waves of COVID 19 between January 2020 and January 2021. The fifth wave caused by the Omicron variant, however, resulted in a total of >1 million cases and >9,000 Covid-19 associated deaths from January 6 to Oct 23, 2022 [5].

Airway epithelium is one of the first infected human tissues by SARS-CoV2. Studies have shown that angiotensin converting enzyme-2 (ACE2) [6] and transmembrane proteases serine 2 (TMPRSS2) [7], which are the main entry factors for SARS-CoV-2, can be identified in human epithelial tissues, including nasal epithelium. Not only do nasal epithelial cells serve as the entry site, but nasal mucosa also acts as the first line of defense against the SARS-CoV-2 entry. While the mucus provides the biochemical barrier, the adaptive immunity on the mucosal surface is equally important to limit the invasion of SARS-CoV-2.

Previously, we measured the SARS-CoV-2 spike 1 (S1)- protein specific antibodies in nasal epithelial lining fluid (NELF) in 81 Covid-19 patients from disease onset to six months after discharge [8] and in 83 unexposed Covid-19 vaccine recipients [9]. The induction of nasal antibody response is different between natural infection and mRNA vaccine. We found that in these Covid-19 patients who had no pre-existing immunity from vaccination before their infection (recruited in the early phase of the pandemic, June 2020 – January 2021), their nasal antibody was IgA dominant with barely detectable IgG. Moreover, the nasal IgA was induced earlier than their plasma counterpart, and being detected as early as on the fourth day post-diagnosis. In addition, the NELF could inhibit the binding of SARS-CoV-2 to ACE2 which infers its neutralizing ability against SARS-CoV-2 *in vivo*. At six months post-diagnosis, half of recovered subjects still possessed S1-specific IgA in their NELF. In contrast, in the unexposed subjects who received mRNA vaccine, S1-specific IgA and IgG were detectable in 40% and 8% of their NELF by 14 ± 2 days after the first dose and 82% and 68% by 7±2 days after the second dose, respectively. Strikingly, the induction of S1-specific antibody was not detected in the unexposed subjects who took inactivated vaccine.

Currently, the mRNA vaccine is widely used in western countries while the inactivated vaccine is available mainly in developing countries. The enhancement of serological antibody response and cellular immunity could be observed after three, or even four doses of either mRNA vaccine or inactivated vaccine [10,11]. However, most of the studies did not report the local immunological parameters. Concurrently, with the progression of the pandemic as well as the availability of vaccine in different formats, our population has also acquired ‘hybrid’ immunity against SARS-CoV-2 from a combination of scenarios, e.g., a natural infection before the availability of vaccine, vaccination after Covid-19 recovery, unexposed with different vaccine regimens, vaccinated but eventually contracted Covid-19, or any of the above with re-infection. As more individuals were infected with Covid-19, it would be of clinical relevance to evaluate the benefit of further doses of vaccine in enhancing the durability, antibody breadth and the neutralizing potential of mucosal and circulating antibody in subjects after recovery from infection. Moreover, as we found that nasal immunity could be induced by current mRNA vaccine or prior Covid-19 infection, it is important to find out if this could boost the mucosal immune response in the recovered patients.

Unlike the previous VOCs, the Omicron variant has thirty-seven mutations in the spike protein, fifteen of which are present in the receptor binding domain (RBD) [12]. These mutations enhanced the binding ability to human ACE2 and weakened the binding ability of the antibodies induced by the non-Omicron SARS-CoV-2 or vaccine designed against the ancestral strain [13]. The reduced neutralizing ability against the Omicron variant were observed in serological study [12]. However, whether nasal immune response to the Omicron variant could be boosted by the current mRNA vaccine (BNT162b2) was not well studied.

This study describes the kinetics of SARS-CoV-2 S1-specific antibody isotypes (IgA and IgG) in the NELF of recovered subjects and vaccine-only subjects from either the day of disease onset or at baseline, i.e., 0-to-2 days before vaccination, to six months of the initial event. The result of this study provides the antibody level, durability and the neutralizing potential in the NELF and plasma against the ancestral and omicron BA.1 strains of SARS-CoV-2. This study will improve our understanding on the antibody isotype kinetics and neutralizing capacity induced among adults with natural infection, vaccination and hybrid immunity.

## 2. Materials and Methods

### 2.1. Subject Recruitment

The study cohort was followed from the early phase of the pandemic (August to December 2020), before the emergence of SARS-CoV-2 VOCs. Adult patients who were hospitalized with Covid-19 were recruited prospectively if they were within four days of their first RT-PCR-positive result [7]. The disease status was confirmed by two RT-PCR tests targeting different regions of the RdRp gene performed by the Public Health Laboratory Service by the Centre of Health Protection. Patients were allocated to the Prince of Wales Hospital in the East New Territories of Hong Kong for clinical management. All patients were unvaccinated and without known prior SARS-CoV-2 infection. Day 0 was considered as the first day of symptoms. Patients were discharged when they were consecutively tested negative for SARS-CoV-2 by RT-PCR or had a viral threshold cycle (CT) value of above 32 and tested positive for nucleocapsid specific serum IgG. All the eleven recovered subjects took one dose of mRNA vaccine (BNT162b2) with at least 180 days interval between infection and vaccination.

Eleven unexposed but vaccinated subjects (control subjects) with similar age (+/-3 years old) and same gender were recruited as the vaccine-only group. These subjects were confirmed with no known SARS-CoV-2 infection by CT value of above 40 at 0-2 days before vaccination and the absence of mucosal and serological antibodies against SARS-CoV-2 S1-protein in their baseline specimens. These subjects took the first and second dose of mRNA vaccine on Day 0 and Day 21. The third dose (booster dose) of mRNA vaccine was taken at least 180 days after the first dose. They reported that they did not experience any SARS-CoV-2 infection within the study duration.

All research subjects provided written consent for enrollment with approval from the Joint Chinese University of Hong Kong—New Territories East Cluster Clinical Research Ethics Committee (CREC: 2020.076, 2020.4421 and 2021.214).

Longitudinal biospecimen collections of the recovered group were conducted at eight time points during the in-patient & recovered period and post-vaccination period, including disease onset (onset), 4 weeks (4W), 13 weeks (13W) and 25 weeks (25W) after onset; 0-to-2 days before vaccination (PreB), 2weeks(B2W), 9weeks(B9W), 21(B21W) after vaccination (**Figure 1A**)

**Figure 1.**
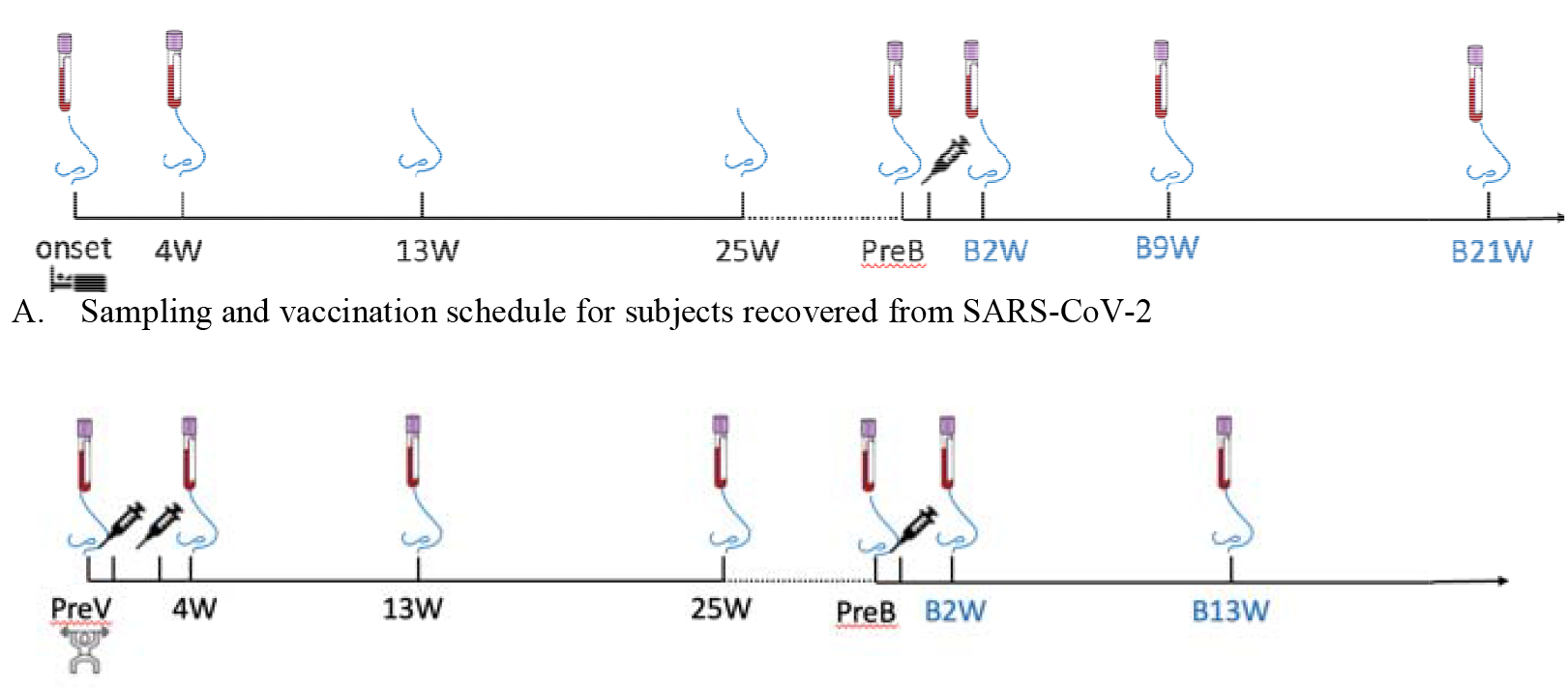
A longitudinal sample collection in (**A**) recovered subjects from the day of diagnosis (disease onset) to twenty-one-week post-vaccination and in (**B**) vaccine-only subjects from 0-to-2 days before the first dose of vaccine to thirteen weeks post-third dose of vaccine. The nose and blood cartoons indicate the time points when NELF and plasma were collected. The syringe cartoon indicates the time points when the subjects received vaccines.

The specimens in vaccine-only group were also collected at seven time points, including 0-to-2 days before the first dose (PreV), 4 weeks (4W), 13weeks (13W) and 25weeks (25W) after the first dose of vaccine, 0-to-2 days before the third dose (PreB), 2 weeks (B2W (**Figure 1B**).

### 2.2. Severity Scoring

Disease severity was categorized as described in the World Health Organization’s Covid-19 clinical management living guidance [14]. The disease severity of the symptomatic subjects was categorized into mild (where the clinical symptoms were light, and there was no sign of pneumonia on imaging), moderate (with fever, respiratory tract problems and other symptoms, with imaging suggesting pneumonia), severe (coinciding with any of the following: (1) respiratory distress, respiration rate (RR) ≥ 30 times/min; (2) oxygen saturation of ≤ 93% in the resting state; (3) PaO_2_ / FiO_2_ ≤ 300 mmHg (1 mmHg = 0.133 kPa)).

### 2.3. NELF Collection

The nasal strip, made of Leukosorb, was inserted into each nostril after 100 μL of sterile saline was instilled followed by a one-minute nose pinch as described [15, 16]. All strips were collected and transferred to a sterile collection tube and eluted within 24 h after collection.

### 2.4. Elution of NELF and the Preparation of Plasma

To elute the NELF, nasal strips were soaked in 300 μL of phosphate-buffered saline (PBS) on ice. The solution and the strips were transferred to a Costar Spin-X (CLS9301) and centrifuged at 4 °C. 3 mL of blood was collected by venipuncture and transferred into an EDTA blood tube. Plasma samples were separated by centrifugation at 4 °C at 2000 g for 20 min. The specimens were aliquoted into small volume vials and stored at −80 °C until the downstream analysis of SARS-CoV-2-specific Ig panels and neutralization tests.

### 2.5. Measurement of SARS-CoV-2 Spike Protein-Specific IgA and IgG

Semi-quantitative measurements of SARS-CoV-2 spike protein (S1 domain)-specific Ig ELISA Kits (Euroimmun, EI 2606-9601 A and EI 2606-9601 G) were used. For this measurement, 1:10 diluted-NELF, as well as 1:100 diluted plasma, were assayed following the manufacturer’s instructions and analyzed with a Synergy HTX Multi-Mode Reader. A semi-quantitative readout was used for the ratio between the sample and the calibrator’s optical density (OD). Data were expressed in the sample/calibrator (S/C) ratio, where a value of ≥ 1.1 was considered positive.

### 2.6. Measurement of SARS-CoV-2 Neutralizing Antibody (NAb) against the ancestral SARS-CoV-2 and omicron BA.1

A blocking enzyme-linked immunosorbent assay (GenScript, L00847) was employed as a surrogate of the neutralization test. Briefly, undiluted NELF, 1:10 and 1:100 diluted plasma samples, and controls were processed as per the manufacturer’s instructions. Samples that gave a signal inhibition of ≥ 30% were considered to be SARS-CoV-2 NAb-positive.

### 2.7. Statistical Analysis

The demographic variables of the subjects were described by medians and range for continuous variables and frequencies and percentages for categorical variables. For the immunoglobulin profile comparisons between the recovered group and the vaccine-only groups were assessed using Wilcoxon matched-pairs signed rank test and Fisher’s exact test, as appropriate. All the S1-specific IgA and IgG levels were expressed as median S/C ratio. All statistical tests were performed using GraphPad version 9.4.1 for the macOS. Differences were considered statistically significant at *p* < 0.05 on a two-tailed test.

## 3. Results

### 3.1. Demographics of the subjects recruited

The cohort consisted of eleven recovered subjects who had participated in an in-patient study [8] and eleven age- and gender-matched seronegative individuals before their first dose of vaccination who had participated in a longitudinal vaccination study since early 2021 [9] (**Table 1**). The median age was 62, ranging from 17-69 years old. Four were male and seven were female. All Covid-19 patients were symptomatic with four mild, four moderate, and three severe cases. The median hospitalization was 14 days, ranging from 9-20 days. The eleven subjects got infected from August 2020 to December 2020. The median duration between onset and one dose of vaccine was 242 days, ranging from 206 days to 311 days. All these eleven subjects took one dose of mRNA vaccine. No death cases were included in this study.

**Table 1.**
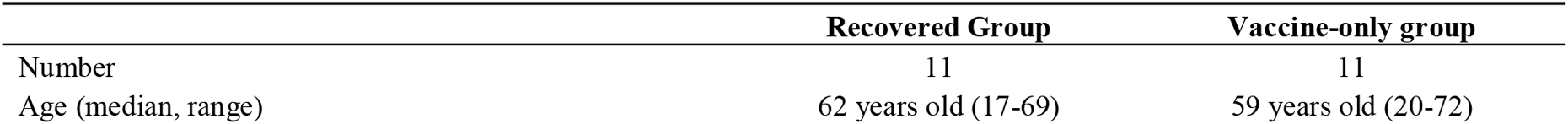
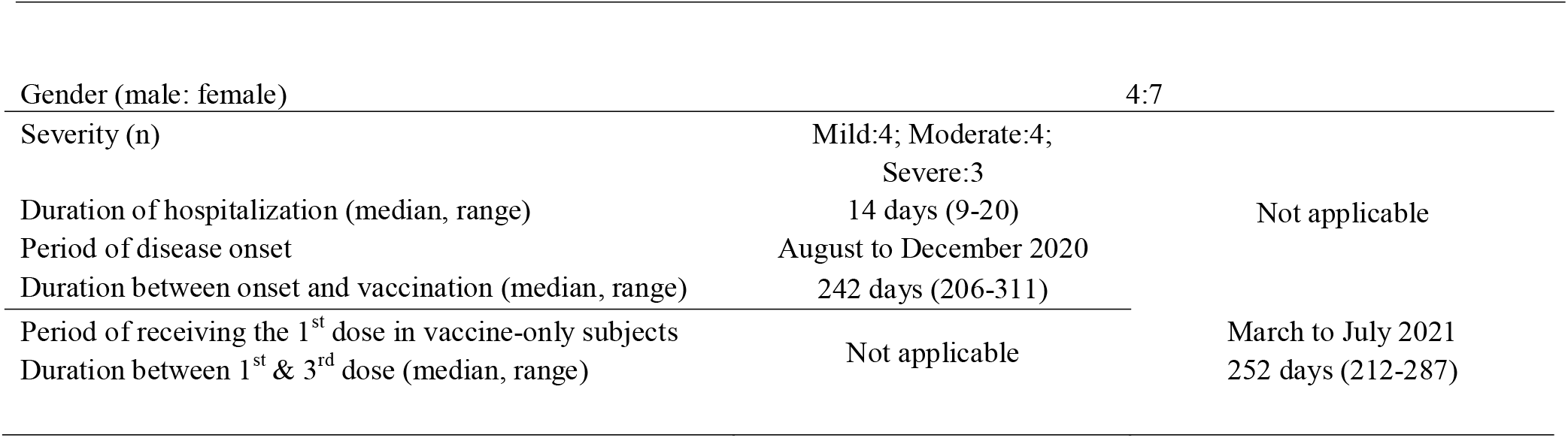
Demographics of the recovered patients and vaccine-only subjects.

### 3.2 SARS-CoV-2 S1 specific antibody levels in NELF and plasma before the booster dose

In the “Recovered Group”, the NELF collected in the fourth week (4W) of disease onset contained S1-specific IgA (S/C ratio = 8.98) but not IgG (S/C ratio = 0.41), and the IgA declined over the 25 weeks post diagnosis but remained detectable (**Figure 2A**). In contrast, a good induction of S1-specific IgA (S/C ratio = 10.87) and IgG (S/C ratio = 6.50) was seen in the plasma of these patients. Though the circulating IgA and IgG declined over the 25 weeks post diagnosis, they remained detectable at the pre-booster (PreB) time point (**Figure 2B**). The S1-antibody longevity was greater in plasma than it was in the nasal cavity.

**Figure 2.**
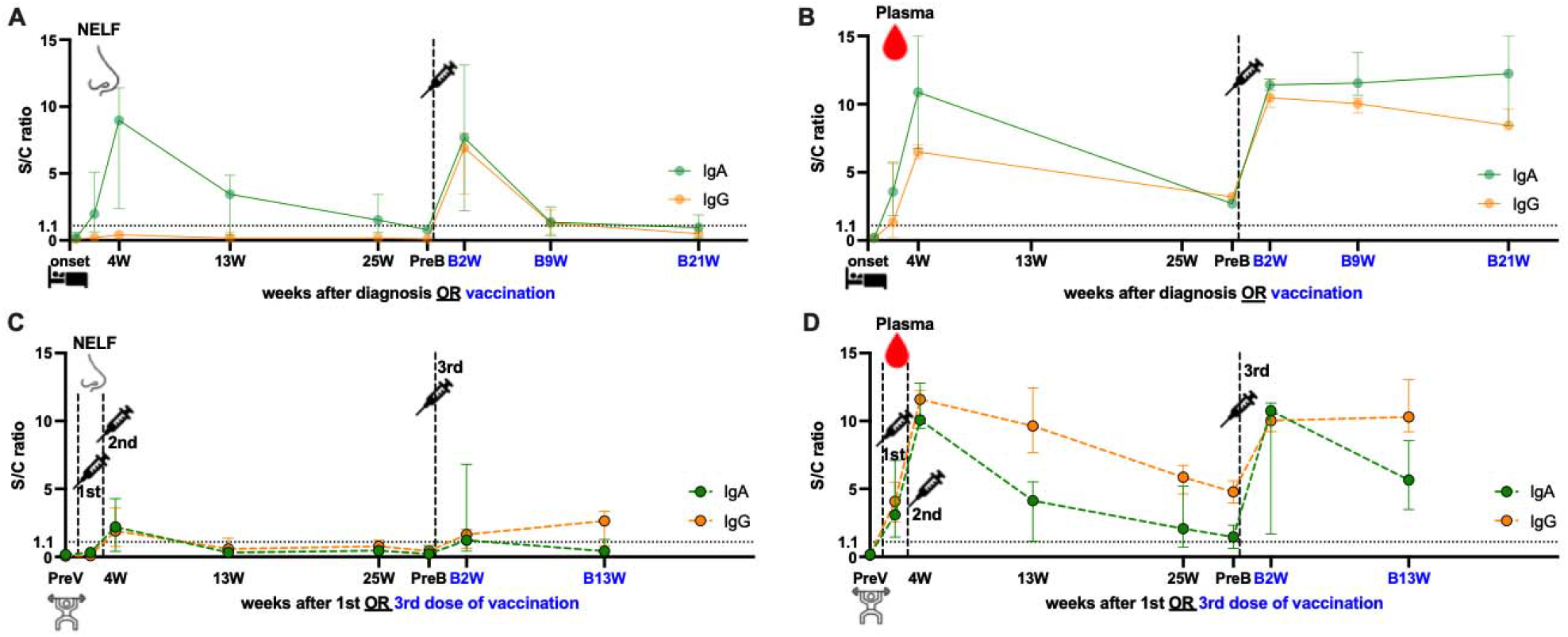
SARS-CoV-2 S1-specific antibody levels in the (A, C) nasal epithelial lining fluid (NELF) and (B, D) plasma of in (A, B) recovered subjects and (C, D) vaccine-only subjects. Antibody-level data points above the dotted line (sample/calibrator (S/C) ratio ≥ 1.1) are considered positive, while the S/C ratio = 15 indicates the upper detection limit of the assay. The median and interquartile range are plotted. Green and orange symbols indicate data of IgA and IgG, respectively. The levels of S1-specific Ig were compared between the recovered group and vaccine-only group by the Mann–Whitney rank test. In the recovered group, S1-specific Igs were measured at the onset of the disease, 4 (4W), 13 (13W), 25 (25W) weeks after onset, 0 to 2 days before booster (PreB) and 2 (B2W), 13 (B13W) and 21 (B21W) weeks after vaccination. In the vaccine-only group, measurements were done on 0 to 2 days before the first dose of vaccine (PreV), 4 (4W), 13 (13W), and 25 (25W) weeks after the first dose, 0 to 2 days before the booster (PreB), and 2 (B2W), 13 (B13W) weeks post booster.

In the vaccine-only subjects with two doses of mRNA vaccines, both S1-specific IgA (S/C ratio = 2.2 in NELF; 10.08 in plasma) and IgG (S/C ratio = 1.91 in NELF; 11.59 in plasma) were detected in the NELF (**Figure 2C**) and plasma (**Figure 2D**) four weeks (4W) after receiving the initial dose. However, the induced S1-specific nasal antibodies declined quickly and became undetectable 13 weeks post first dose while the plasma antibodies lasted at least 25 weeks post first dose and remained detectable at the pre-booster (PreB) time point.

### 3.3 The booster dose of mRNA vaccine induced IgG occurence in the NELF of recovered subjects

While the natural infection did not induce any nasal IgG, one dose of mRNA vaccine could expand the S1-specific immunoglobulin isotype in the NELF with both IgA (S/C ratio = 7.68) and IgG (S/C ratio = 6.92) two weeks after receiving booster (**Figure 2A**, B2W). Nevertheless, both nasal IgA and IgG dropped quickly and became marginally detectable at nine weeks post booster (B9W). In contrast, the circulating IgA and IgG were boosted with a greater magnitude and remained at a high level for at least 21 weeks after the booster dose (**Figure 2B**, B21W). Moreover, the levels of IgG detected at B21W were even higher than the convalesce phase (4W) (IgG: S/C ratio = 8.44 at B21W, 6.50 at 4W, *p* = 0.0364), while IgA also got a similar trend (S/C ratio = 12.25 at B21W, 10.87 at 4W, *p* = 0.7091).

Unlike the response in the recovered subjects, the booster dose did not induce extensive production of nasal IgA and IgG in the “vaccine-only” subjects who had received two doses of mRNA vaccines. The nasal S1-specific IgA (S/C ratio = 1.23 at B2W) and IgG (S/C ratio = 1.64 at B2W) showed no statistical difference from those induced at 4W. The low level of nasal IgA gradually decreased and became below the positive cut-off in the 13^th^ week of booster (**Figure 2C**, B13W). Nasal IgG lasted subtly longer than IgA and remained positive at B13W. Nevertheless, the booster dose induced the production of plasma S1-specific IgA (S/C ratio = 10.75 at B2W) and IgG (S/C ratio = 10.03 at B2W) from their PreB level, however, they were not exceeding the highest level found at four weeks after the first dose (4W). Still, both plasma S1-IgA and IgG remained at high levels 13 weeks post booster, though plasma IgA waned quicker than IgG (**Figure 2D**, B13W).

To summarize, the booster dose induced higher of nasal IgA and IgG, and plasma IgA in recovered subjects than vaccine-only subjects. We found that the S/C ratio of nasal specific IgA (7.68 vs 0.42, *p* = 0.0115), IgG (6.92 vs 2.65, *p* = 0.0190), and plasma IgA (11.43 vs 5.66, *p* = 0.0004) in the recovered group were significantly higher than that in the vaccine-only group while plasma IgG levels were similar in the two groups (10.48 vs 10.29) (**Supplementary Figure 1**).

### 3.4 The plasma of recovered patients exerted a stronger inhibition against the binding of ACE2 to the ancestral SARS-CoV-2 than the vaccine-only group

In Figure 3A, we showed that in the recovered group, 8/11 NELF (open circles) and 9/11 plasma (open squares) samples contained NAb against the ancestral RBD at four weeks after onset (4W). The booster dose did not increase the proportion of NAb positive NELF (7/11, black dots) but enriched all recovered subjects’ plasma with NAb (11/11, black squares). In contrast, only 7/11 and 4/9 of the vaccine-only subjects had NAb in their NELF at time points V2 and V3 (blue dots), respectively, while all plasma samples contained NAb against ancestral SARS-CoV-2 RBD at both time points (blue squares). This infers the high potency of mRNA vaccine in inducing circulating NAb.

**Figure 3.**
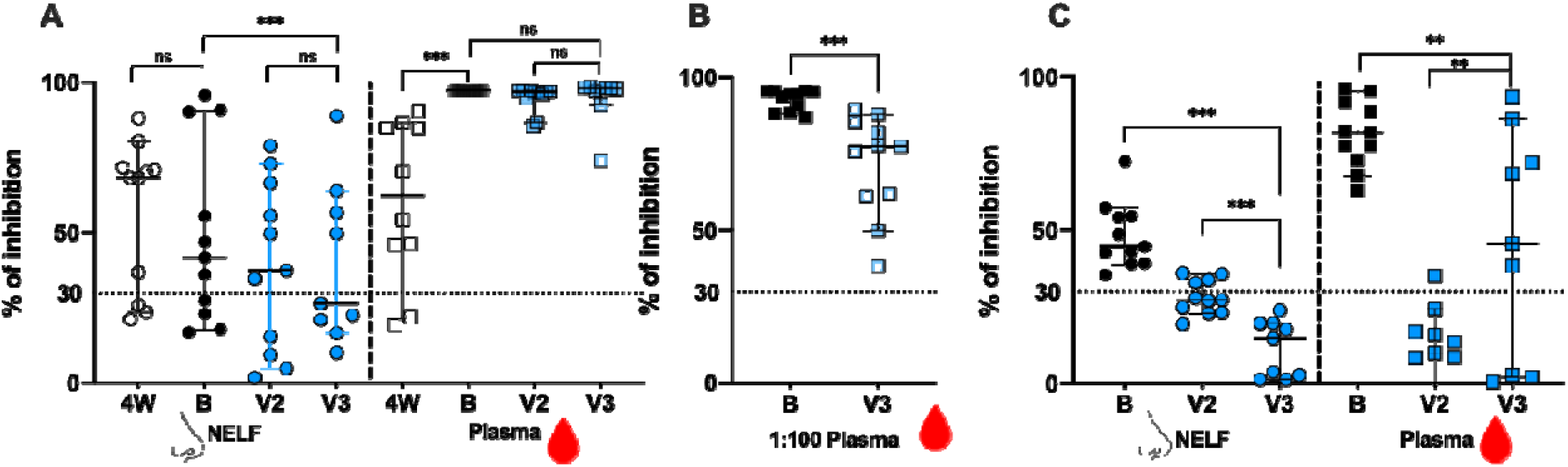
The signal inhibition in the surrogate ACE-2-based neutralization readout. (A) The percentage of signal inhibition against ancestra RBD of SARS-CoV-2 by NELF (circle) and plasma (square) of recovered patients (4W: 4 weeks after onset; B: 2 weeks post booster) and vaccine-only subjects (V2: 4 weeks post 1^st^ dose; V3: 2 weeks post 3^rd^ dose) are plotted. (B) Plasma diluted at 1:100 dilution was used to provide a better resolution to examine the differential neutralization ability between recovered (black square) and control (blue square) subjects. (C) The percentage of signal inhibition against SARS-CoV-2 omicron BA.1 by NELF and plasma of recovered patients after booster (B: 2 weeks post vaccination) and vaccine-only subjects (V2: 4 weeks post 1^st^ dose; V3: 2 weeks post 3^rd^ dose) are plotted. The ≥ 30% signal inhibition cutoff for SARS-CoV-2 NAb detection is interpreted as the sample containing neutralizing antibodies for SARS-CoV-2, indicated by the horizontal dotted line. The median and the 95% CI are shown. Comparison was performed with a Wilcoxon matched-pairs rank test. The asterisks indicate the statistical differences found; *: *p* < 0.05, **: *p* < 0.01 and ***: *p* < 0.005.

When the inhibition competence was compared quantitatively before and after booster, no enhancement of NAb against the ancestral SARS-CoV-2 was found in the NELFs of both recovered patients (68.22% at 4W vs 41.65% at B, *p* = 0.3203) and of the vaccine-only group from V2 to V3 (37.4% at V2 and 26.4% at V3, *p* = 0.1289). The booster was only effective in increasing the NAb in the plasma of the recovered group (62.30% at 4W vs 97.31% at B, *p* = 0.0020) while those in the vaccine-only group remained above 96% (96.75% at V2 vs 97.89% at V3, *p* = 0.1289, **Figure 3A**).

When comparing the NAb between the recovered and vaccine-only groups after booster, the recovered group had a significantly stronger inhibition effect in their NELF (41.65% vs 26.4%, *p* = 0.0039) and plasma. As a high number of plasma samples gave a saturated readout, the plasma samples were diluted in 1:100 for further evaluation. In Figure 3B, the 1:100 plasma of the recovered group had a significantly higher percentage of binding inhibition effect than that in the vaccine-only group (94.61% vs 77.42%, *p* = 0.0010).

### 3.5 Booster provided a stronger inhibition against the omicron BA.1 variant in the recovered group

As we did not have enough volume of NELF and plasma sample at four weeks after onset for evaluation of their binding inhibition efficacy towards omicron BA.1, we could only report the comparison between the recovered group and vaccine-only group after booster and the effect of booster within the vaccine-only group (**Figure 3C**). The booster provided all recovered subjects with NAb against the omicron BA.1 in their NELF (44.69%, black dots) and plasma (81.81%, black squares).

Surprisingly, four vaccine-only subjects were found to have NAb against omicron BA.1 before their booster (V2) while the booster could not induce detectable NAb against omicron at V3 (14.68%) which was similar to its effect against ancestral SARS-CoV-2 in the NELF. Intriguingly, the booster was still effective in enhancing the proportion of vaccine-only subjects from 1/11 at V2 to 6/9 at V3 in having the NAb against omicron BA.1. The inhibition efficacy in the plasma of the vaccine-only group rose from below detection limit (9.87%) at V2 to 45.60% at V3 (*p* = 0.0117). Finally, the levels of NAb against omicron BA.1 RBD in the NELF *(p* = 0.0039) and plasma (*p* = 0.0117) of the recovered group were significantly higher than those in the vaccine-only group.

## 4. Discussion

Understanding the mucosal antibody dynamics is an important aspect of evaluating the protection induced by natural infection and any vaccine candidates, as it is one of the keys to sterilizing immunity [17,18]. In animal models, antibodies alone are sufficient to protect against SARS-CoV-2 infection [19]. By stimulating mononuclear cells isolated from the tonsil of SARS-CoV-2 infected individuals, Mahallawi et al. confirmed that the SARS-CoV-2 spike primed potent specific memory B cells in nasal associated lymphoid tissue (NALT) [20]. The mRNA vaccine, which induces nasal antibody in unexposed subjects, may have the potential to induce the recall of NALT specific memory B cells, thus the increase in nasal Ig levels in the NELF of the recovered subjects receiving their booster. However, no study reported whether the mRNA vaccine alone could induce specific memory B cells in NALT. Tang et al collected immune cells in bronchoalveolar lavage fluid (BAL) from the Covid-19 infected subjects and vaccine-only subjects. They found that 0.25-8% of total B cells in the infected subjects were RBD^+^ B cells whilst the vaccine-only subjects had a lower percentage at around 0-1% [21]. The lack of local RBD^+^ B cells in vaccine-only subjects may explain why the third dose of mRNA vaccine did not boost mucosal Ig extensively in the vaccine-only group. Besides, we noticed the change of nasal antibody isotypes by mRNA vaccine in the recovered groups, with a significant rise of nasal specific IgG while it was negative before the booster. Nasal IgA and IgG induced by the booster lasted only 13 weeks, which is shorter than that acquired after natural infection.

We observed a significant increase of plasma NAb against the RBD of the ancestral strain after one dose of vaccine in the recovered subjects, especially in two of them who had negative NAb before vaccination. The available data in the literature indicated that vaccine-only subjects had weaker neutralizing serum responses, as half of their RBD-specific memory B cells displayed high affinity toward multiple VOCs, while the boosted recovered subjects had their memory B cell pool expanded selectively, matured further and harbored more mutations in their variable V_H_ genes [22]. This could explain the boosted neutralizing ability against the ancestral and the omicron BA.1 in the plasma of the recovered group. Interestingly, the nasal neutralizing ability against the ancestral virus induced by mRNA vaccine in the recovered group was much stronger than the vaccine-only group. As the specific memory B cells response in NALT is poorly studied in mRNA vaccinees, no direct evidence illustrates the correlation of memory B cell response and the strong NAb in nasal mucosa.

We found that seven subjects with positive nasal NAb after infection continued to have positive NAb after mRNA vaccine while the NAb in another three subjects remained negative after infection and vaccination. A similar result was observed in the vaccine-only subjects after two and three doses of vaccine **(Figure 3A**). This infers that some subjects might have impaired nasal immune response so that either there were no inductions at the lamina propria, or intrinsic IgA deficiency [23], or the IgA produced did not undergo transcytosis by the polymeric immunoglobulin receptor (pIgR) and therefore, no secretory IgA was detected in the NELF of these subjects [27].

More importantly, all eleven recovered subjects acquired nasal neutralizing antibodies against omicron BA.1 variant after one dose of mRNA vaccine, and their median binding inhibition was significantly stronger than the vaccine-only group. As we lacked nasal NAb data against omicron BA.1 directly after natural infection and there was no published data evaluating the same aspect, it is not clear if the booster was potentiating the inherited effect from the prior natural infection, or it was expanding the antibody breadth. Nevertheless, some studies reported that NAb against the omicron BA.1 variant was detectable but weak in other mucosal fluids, e.g., saliva [24] and BAL [21], after non-omicron Covid-19 infection. In particular, Diem et al. reported that the saliva of the non-infected subjects after three doses of mRNA vaccine had a comparable neutralizing titer against Delta, BA.1, and BA.2 to those recovered subjects [24]. In a similar scope, our research suggested that one dose of mRNA vaccine could induce positive nasal NAb in Covid-19 recovered subjects. Together with the better serological Nab, the booster would provide a better immune protection against the omicron variant.

It is noteworthy that three doses of mRNA vaccine did not boost the nasal NAb against the ancestral SARS-CoV-2 nor omicron BA.1 variant in the vaccine-only group, although positive nasal Ig were detected after mRNA vaccine. This could be the reason why mRNA vaccine could not provide sterile protection to its vaccinees. In contrast, a significant increase of plasma NAb against the omicron variant was detected after the third dose, which is consistent with other studies [25,26] and contributed to its protection against disease severity. These results suggested the intrinsic difference in the induction and potentiation of local and circulating antibody. Therefore, internasal vaccines are now under preclinical research and some of them could produce protection against SARS-CoV-2 in the upper and lower respiratory tract in animal models [28-30]. All in all, the B cell response elicited by the mRNA and other vaccine candidates in NALT and mucosal sites deserve a full examination for better vaccine design.

Apart from humoral immunity, cellular immunity is also critical to combat viral infections. Goel et al. reported that mRNA vaccination generated antigen-specific CD8+ T cells and durable memory CD4+ T cells in SARS-CoV-2 naïve and recovered subjects. They also observed an increasing and fast antibody responses to mRNA vaccine in the short-term without significantly altering antibody decay in the recovered subjects [31]. The mucosal-associated T cell response to mRNA vaccine is controversial. One research found that two doses of mRNA vaccine could induce nasal tissue-resident memory (Trm) CD8^+^ T cells in healthy donors, inferring that nasal T cells may be induced and contribute to the protective immunity afforded by this vaccine [32]. Another research reported that mRNA vaccine did not elicit strong S-specific CD8^+^ or CD4^+^ T cell responses in the BAL of SARS-CoV-2 naïve subject while BAL from Covid-19 convalescents had higher cytokine-producing CD8^+^ and CD4 T^+^ cells, indicating that mRNA vaccine may offer limited protection against breakthrough infection [21]. Further studies in our group would focus on the cellular immune response in these two groups of subjects, especially at the mucosal sites.

Meanwhile, there are several limitations in the current study in terms of the generalizability. First, we had a very small sample size, as research subjects with SARS-CoV-2 exposure before vaccination or vaccination without any SARS-CoV-2 exposure were difficult to recruit. The initial pool of Covid-19 patients during the study period was small due to the unique infectious control measures in Hong Kong. Very soon after the first wave, citizens were provided with Covid-19 vaccine from different vendors, including Comirnaty (mRNA vaccine) and CoronaVac (inactivated vaccine). Therefore, the number of Covid-19 cases without prior vaccination became even less available. Although the number of subjects is small, the overlapping between groups was small with their unique pattern. Therefore, without being able to include more subjects, the current pattern is robust to describe the overall pattern. Second, we did not examine the cellular immunity, e.g., lung-resident memory T cells in these subjects, which is another essential arm of immunity to protect us from the next infection. Third, we only evaluated the IgA and IgG dynamics of the S1-specific antibody but not the antibody against other S ectodomains, e.g., the most potent neutralizers against RBD-2 and the greatest recognition breadth S2-1, and viral proteins. Fourth, due to shortage of sample volume, we were not able to determine if the nasal antibody of the recovered patients exhibited higher cross-neutralization breadth than those induced in unexposed vaccine recipients before their boosters. Lastly, we attempted to recruit patients and unexposed subjects who took inactivated vaccine instead of the mRNA vaccine to provide extra information for patients recovered from covid-19 to study the response to vaccines with different mechanisms of action. However, we only recruited two within the study period and cannot provide an explicit picture for discussion within this manuscript. Nevertheless, we want to emphasise that mucosal antibody response is an understudied area because of its difficulties in sample collection and standardization for reliable comparisons. The value of our study is obvious because of the eight consecutive longitudinal sample collections together with the long follow up period. Our research provided the dynamics of antibody changes in nasal fluid and plasma with a sampling period covering two years since disease onset.

## 5. Conclusions

In our study, the “hybrid” immune model (infection followed by mRNA vaccine) induced better nasal antibodies, as well as NAb against the ancestral SARS-CoV-2 and omicron BA.1 variant than the vaccine-only subjects. In circulation, both “hybrid” immune model and vaccine-only group demonstrated boosted antibody response. Our findings suggested that one dose of mRNA vaccine is necessary to maintain the plasma NAb against the ancestral SARS-CoV-2 and elicited the NAb against omicron BA.1 variant for recovered subjects during the omicron wave. The third dose would provide extra benefit for people who had no prior SARS-CoV-2 exposure to acquire serological NAb against SARS-CoV-2 VOC, e.g., omicron BA.1. Further studies focusing on the cellular immunity at the mucosal sites will be needed to elucidate the comprehensive outcome of the hybrid immunity. Finally, though the differential pattern between the hybrid and vaccine-only group is robust, cautions should be taken for its generalizability due to its unavoidable small sample size.

## Data Availability

All data produced in the present work are contained in the manuscript

---

**Supplementary Figure 1.**
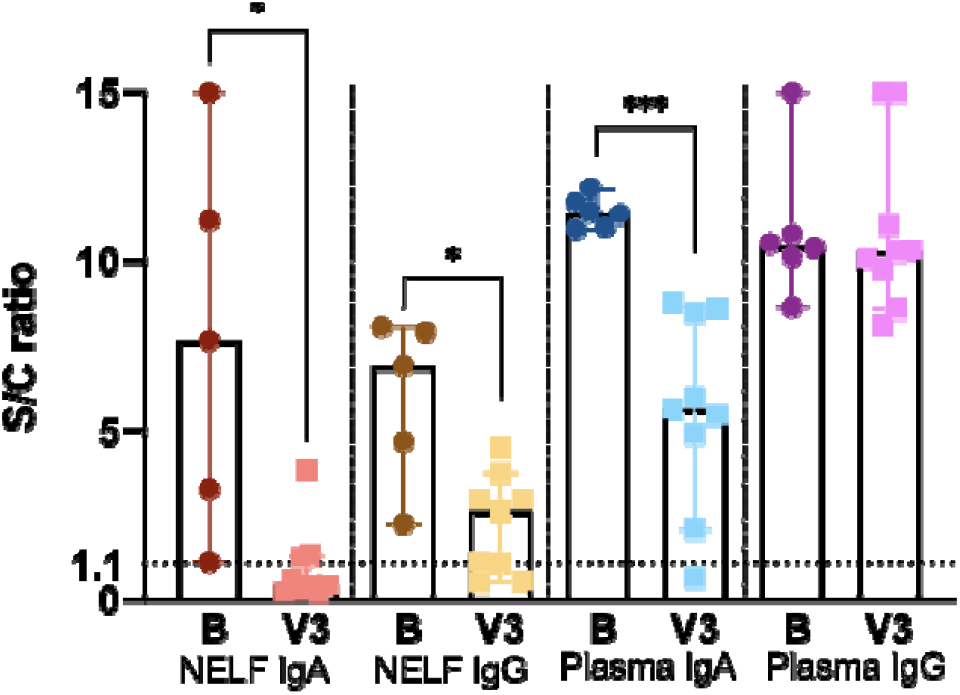
SARS-CoV-2 S1-specific Ig in the recovered group (dots) and the vaccine-only group (squares) at two weeks post booster. B: 2 weeks post mRNA vaccine in the recovered group; V3: 2 weeks post third dose in the vaccine-only group. The levels of S1-specific Ig were compared between groups using Mann-Whitney test. The asterisks indicate the statistical differences found; *: *p* < 0.05 and ***: *p* < 0.005.

## Author Contributions

For research articles with several authors, a short paragraph specifying their individual contributions must be provided. The following statements should be used “Conceptualization, formal analysis, S.L. and R.W.Y.C.; investigation, methodology, S.L., J.G.S.T. and R.W.Y.C.; patient recruitment, S.L., J.G.S.T. and G.C.Y.L.; resources, G.C.Y.L., P.K.S.C. and R.W.Y.C.; writing—original draft preparation, S.L. and R.W.Y.C.; writing— S.L., J.G.S.T., G.P.G.F., G.C.Y.L., K.Y.Y.C.; P.K.S.C., R.W.Y.C.; visualization, S.L. and R.W.Y.C.; supervision, R.W.Y.C.; project administration, S.L., J.G.S.T., R.W.Y.C.; funding acquisition, R.W.Y.C. All authors have read and agreed to the published version of the manuscript.”

## Funding

This work was supported by the Innovation and Technology Fund PRP/039/21FX (R.W.Y.C.), Health and Medical Research Fund-commissioned grants COVID190112 (R.W.Y.C.); and Health and Medical Research Fund 19200131 (R.W.Y.C.).

## Institutional Review Board Statement

The study was conducted in accordance with the Declaration of Helsinki and approved by the Joint Chinese University of Hong Kong—New Territories East Cluster Clinical Research Ethics Committee (CREC: 2020.076, 2020.442 and 2021.214).

## Informed Consent Statement

Informed consent was obtained from all subjects involved in the study.

## Data Availability Statement

The datasets generated are available from the corresponding author on reasonable request.

## Acknowledgments

We would like to acknowledge Chris KP Mok (JC School of Public Health and Primary Care, The Chinese University of Hong Kong) for providing the omicron BA.1 RBD for the surrogate neutralization assay; Aaron HP Ho, Megan YP Ho and Yuan-yuan Wei (Department of Biomedical Engineering, Faculty of Engineering, the Chinese University of Hong Kong) who tailor-made the nasal strips for this study; Dr. Genevieve Fung (Department of Paediatrics, The Chinese University of Hong Kong) for the sample collection, and Fiona Cheng (Department of Paediatrics, The Chinese University of Hong Kong) for her assistance in preparing all the nasal strip vials. We would like to offer our special thanks to Vickie Li (Department of Medicine and Therapeutics, the Chinese University of Hong Kong), Apple Yeung and Lam Lap Yee (Department of Microbiology, the Chinese University of Hong Kong) for their assistance in the sample logistics. We thank all the subjects who agreed to participate in this study.

## Conflicts of Interest

The authors declare no conflict of interest.

## Notes

### Competing Interest Statement

The authors have declared no competing interest.

### Funding Statement

This study was funded by the Innovation and Technology Fund PRP/039/21FX (R.W.Y.C.), Health and Medical Research Fund-commissioned grants COVID190112 (R.W.Y.C.); and Health and Medical Research Fund 19200131 (R.W.Y.C.).

### Author Declarations

The Joint Chinese University of Hong Kong New Territories East Cluster Clinical Research Ethics Committee gave ethical approval for this work.

